# Post-covid medical complaints after SARS-CoV-2 Omicron vs Delta variants - a prospective cohort study

**DOI:** 10.1101/2022.05.23.22275445

**Authors:** Karin Magnusson, Doris Tove Kristoffersen, Andrea Dell’Isola, Ali Kiadaliri, Aleksandra Turkiewicz, Jos Runhaar, Sita Bierma-Zeinstra, Martin Englund, Per Minor Magnus, Jonas Minet Kinge

**Affiliations:** Norwegian Institute of Public Health, Oslo, Norway; Clinical Epidemiology Unit, Orthopedics, Department of Clinical Sciences Lund, Lund University, Lund, Sweden; Centre for Economic Demography, Lund University, Lund, Sweden; Department of General Practice, Erasmus MC University Medical Center Rotterdam, Rotterdam, the Netherlands; Department of Orthopedics & Sports Medicine, Erasmus MC University Medical Center Rotterdam, Rotterdam, the Netherlands; Department of Health Management and Health Economics, University of Oslo, Oslo, Norway

**Author notes:** **Corresponding author, contact information:** Karin Magnusson, Cluster for Health Services Research, Norwegian Institute of Public Health, Postboks 222, Skøyen, N-0213 Oslo. Visiting address: Sandakerveien 24c, Building D, 0473 Oslo.

## Abstract

**Objective:** To examine whether persons infected with the SARS-CoV-2 omicron variant have an altered risk of post-covid complaints and healthcare use when compared to 1) persons testing negative, and 2) persons with delta.

**Design:** Prospective cohort study with 126 days follow-up and a time-to-event approach.

**Setting:** A registry-based study including Norwegian residents.

**Participants:** All persons aged 18-70 years living in Norway and who had a negative polymerase chain reaction (PCR) test for SARS-CoV-2 (N=105 196, mean (SD) age 42 (14), 50% women)) or positive test with confirmed omicron variant (N=13 028, mean (SD) age 37 (13), 50% women) or delta variant (N=23 368, mean (SD) age 40 (12), 50% women) in December 2021. Individuals with hospital contacts or non-screened PCR test were excluded.

**Main outcome measures:** Symptoms/complaints and diagnosis of musculoskeletal pain, fatigue, cough, heart palpitations, shortness of breath, anxiety/depression and brain fog at the general practitioner or emergency ward as recorded in national registries and as observed for the whole follow-up period as well as in periods 14-30 days, 30-90 days and 90 days or more.

**Results:** Persons with omicron or delta had similarly increased risk of post-covid fatigue compared to persons testing negative, with a hazard ratio (HR)=1.21 (CI:1.10-1.33) for omicron and HR=1.26 (CI: 1.17-1.35) for delta, for up to 126 days after the test date. They also had an increased risk of shortness of breath (HR=1.43, CI, 1.14-1.80 and HR=1.70, CI, 1.46-1.98 for omicron and delta, respectively, relative to negative). Omicron was related with a similar, and no increased risk of musculoskeletal pain, cough, heart palpitations, anxiety/depression when compared to delta and when compared to test negative. The risk of complaints was similar for omicron and delta and decreased over time for the post-covid periods 14-30 days, 30-90 days and 90 days or more.

**Conclusions:** SARS-CoV-2 omicron and delta infection are associated with similarly increased risks of post-covid complaints when compared to non-infected. The omicron variant will likely lead to a temporarily increased burden on healthcare services.

## Introduction

An increase in medical complaints following mild SARS-CoV-2 infection, sometimes referred to as “long-covid”, has been reported^1,2,3^. Although the SARS-CoV-2 omicron variant has been associated with less severe acute disease and a reduced risk of hospitalizations compared with delta, concerns remain as to whether long-term complaints persist to a similar extent as for earlier variants ^4,5^.

Given the increased secondary attack rate when the index case has omicron rather than delta^6^, and the expectancy of many symptomatic but less serious cases even among vaccinated individuals^7^, there is a need for knowledge of post-omicron risks for medical doctors, health personnel and health policy makers. If the omicron variant leads to temporary or persistent post-covid complaints, it may impose a large burden on health care and society.

Survey data has been used to determine patterns of symptom persistence following SARS-CoV-2 infection, however, the estimates vary extensively, and they cannot be used to infer on the consequences for the healthcare services. For example, dyspnoea after recovery from primary SARS-CoV-2 infection has been reported in 10–20% of patients in one survey study and up to 75% of patients in another survey study^8,9^. Reporting and response biases will affect the accuracy of both symptoms and testing, leading to questionable validity and difficulties with comparisons between studies. Nordic National register data is based on healthcare services that are freely available for all inhabitants, i.e. a medical record as seen in primary care represents both an indication of a complaint as experienced by the patient (and judged so by the medical doctor), and it represents a healthcare contact placing a certain demand on the healthcare services.

The linkage of such medical record data to data on variant-specific SARS-CoV-2 infection including the novel omicron variant can provide unique insights into both post-covid etiology and the expected burden on healthcare systems when many are vaccinated and have mild disease courses. Thus, for the first time, we have studied whether persons infected with the omicron variant have an altered risk of post-covid complaints compared with 1) persons testing negative, and 2) persons with the delta variant. We also provide estimates of prevalent complaints for the acute, sub-acute and chronic post-covid phases, including data beyond 3 months after positive test.

## Methods

### Design and data sources

Using a prospective cohort study design applied to data in the Norwegian Emergency Preparedness Register (S-Table 1),^10^ we included all Norwegian residents aged 18-70 years, who tested negative or positive for SARS-CoV-2 with known variant during the period that the omicron and delta variants had the greatest overlap in Norway; from December 8^th^ to December 31^st^ 2021. These data were linked on the personal ID number to provide information on healthcare contacts in primary care (general practitioners and emergency wards) with specific medical record (S-Table 1). The PCR testing criteria were constant throughout the study period and included persons with symptoms of COVID-19, persons in close contact with anyone with COVID-19 as well as persons having a positive antigen test. Screening for SARS-CoV-2 variant was performed by sanger or whole genome sequencing on all positive PCR tests if the laboratories had capacity and only on positive tests with suspected omicron if the laboratory had capacity challenges. We excluded all persons with unscreened tests and all persons who had a hospital contact from -2 to +14 days from the test date^8^. In this way, we could study persons with known infection with SARS-CoV-2 omicron and delta variant with assumed mild disease courses and/or who were known not to be tested as part of hospital contact or routine testing at hospitals. Participants were categorized into three study groups: 1) persons with omicron, 2) persons with delta, and 3) persons who were non-infected (tested negative). Institutional board review was conducted, and The Ethics Committee of South-East Norway confirmed (June 4th 2020, #153204) that external ethical board review was not required.

### Outcomes

The outcomes included were the most frequently reported post-covid complaints in systematic reviews^11^ (Table 1), as registered in medical records with high validity and reliability^12^ from day 14 after positive test, and onwards: musculoskeletal pain, fatigue, cough, heart palpitations, shortness of breath, anxiety/depression, and brain fog. We allowed for having multiple complaints. If an individual had multiple records with the same complaint within the follow-up period of interest (or combination of diagnostic codes indicative of the complaint, as categorized in Table 1), we chose the first one. At least one day of follow-up was required and observations were censored at day 126, date of death or emigration, whichever came first.

**Table 1.** Condition/complaint with corresponding diagnostic code (ICPC-2) used in primary care.

|  |  |  |
| --- | --- | --- |
|  | Pain (general/multisite and localized pain and symptoms from the musculoskeletal system, not classified elsewhere (neck, back, arms/hands, feet/legs)) | A01, L01-L17, L18-L20, L29 |
|  | Fatigue | A04, A05, A28, A29 |
|  | Cough | R05 |
|  | Heart palpitations | K04, K05, K29 |
|  | Shortness of breath | R02 |
|  | Anxiety and depression | P03, P76, P01, P74 |
|  | Brain fog (concentration or memory problems) | P20 |
| *With condition/complaint we refer to all information that may be included in an ICPC-2 (International Classification of Primary Care 2) code: Diseases, disorders, signs, symptoms, and/or complaints as classified by the physician consulted. |  |  |

### Statistical analyses

First, we calculated the person-time with the number of failures (the outcome in question) and incidence rate with 95% confidence interval for all study groups and all outcomes. Second, we estimated the hazard ratio (HR) with 95% confidence intervals (CI) for having the potential post-covid related complaints/diagnoses in primary care, from 14 to up to 126 days after the test date using Cox regression analyses unadjusted and adjusted for age, sex, education level (no education to >1 year college/university education in four categories), the number of comorbidities in 2020-21 (0-1 vs 2 or more)^13^, the number of previous negative tests in 2020-21 (0, 1, 2 or 3 more) and the number of previous all-cause primary care visits in 2020-21 (0 to 10 or more) as potential confounders. We checked specifically for potential confounding by vaccination status (the number of Covid-19 vaccine doses: 0, 1, 2 or 3 or more). All data on potential confounders were identified in the same data sources, i.e. in National register data covering the entire Norwegian population from January 1^st^ 2020 (only education status was registered in 2019) (S-Table 1).

The delta variant dominated earlier than omicron (S-Figure 1), thus we expected differences in follow-up time by variant. This was handled by stratification on the test week in the regression models. After stratification, visual inspection of Schoenfeld Residuals suggested no violation of the proportional hazard assumption. We also ran sensitivity analyses using conditional logit models matched on test week (the latter unadjusted and with similar adjustments as in our main analyses). In this way, i.e. by choosing the time period where the overlap between the two variants was the greatest, and by having two analytic approaches that match persons infected and non-infected on their calendar week of testing and on their follow-up time, respectively, we could limit any potential bias arising from potentially differential temporal trends in test and screening patterns by variant.

Further, to assess whether the post-covid complaints were more or less common in certain periods after positive test, we repeated the main analyses stratifying the outcome on occurring in the acute phase (14 to 30 days), the sub-acute phase (30 to 90 days) and the assumed chronic post-covid condition phase (90 to 126 days) as recommended in previous studies,^14,15^ i.e. using Cox regression analyses with adjustment for all the described confounders and stratified on the calendar week of testing as described above. In these analyses, the time to event started on the first day of the respective periods, implying that we omitted all outcome observations occurring prior to this day 0. We also censored outcome observations occurring after the last day of the respective period. Thus, an individual could have outcome data in multiple time periods, and for each period and outcome, we studied the time until the first record.

Finally, because persons testing negative may be more prone to get tested and subsequently visit primary care due to (persistent) symptoms from similar bodily systems as those affected by SARS-CoV-2, we repeated the time-differentiated analyses using a comparison group consisting of untested persons (aged 18-70 years, non-hospitalized, never tested for SARS-CoV-2 and assigned a random, hypothetical test date during our study period) in a sensitivity analysis. All analyses were run in STATA MP v. 17.

### Patient and public involvement

No patients were involved in setting the research question, study design, outcome measures, or the conduct of the study. The study was based on deidentified data from Norwegian national registries.

## Results

Of in total 3 656 064 persons eligible for the study, 105 196 persons tested negative during our study period, and 55 853 persons had a positive test result that was screened for SARS-CoV-2 variant (S-Figure 2). Persons with the omicron variant (N=13 028) were generally younger, had higher education, fewer comorbidities and were more often vaccinated than persons with the delta variant (N=23 368) (Table 2). There were also some group differences in the length of follow-up time by study group (Table 3). The mortality during follow-up was low (0.08% (95% CI=0.06-0.10), 0.03% (95% CI=0.01-0.06) and 0.04% (95% CI=0.00-0.06) for persons testing negative or positive with delta and omicron, respectively, with confidence intervals calculated based on Wilson).

**Table 2.** Descriptive characteristics.

|  | Testing negative<br>for SARS-CoV-2 | SARS-CoV-2<br>delta variant | SARS-CoV-2<br>omicron variant |
| --- | --- | --- | --- |
|  | N=105 196 | N=23 368 | N=13 028 |
| Age, mean (SD) | 42.0 (14.1) | 40.1 (12.1) | 36.9 (13.1) |
| Women, n (%) | 52 950 (50.3) | 11 559 (49.5) | 6443 (49.5) |
| Primary school<sup>1</sup>, n (%) | 19 441 (18.6) | 4836 (20.8) | 2606 (20.1) |
| Upper secondary school<sup>1</sup>, n (%) | 36 833 (35.2) | 7819 (33.6) | 4156 (32.1) |
| College/university<sup>1</sup>, n (%) | 41 734 (39.8) | 8914 (38.2) | 5406 (41.7) |
| Born in Norway, n (%) | 80 095 (76.1) | 15 859 (67.9) | 9246 (71.0) |
| ≥2 comorbidities<sup>2</sup>, n (%) | 13 273 (12.6) | 2618 (11.2) | 1149 (8.8) |
| One vaccine dose<sup>3</sup>, n (%) | 97 574 (92.8) | 18 479 (79.1) | 12 096 (92.8) |
| Two vaccine doses<sup>3</sup>, n (%) | 95 487 (90.8) | 17 701 (75.7) | 11 779 (90.4) |
| Three vaccine doses<sup>3</sup>, n (%) | 18 819 (17.9) | 1466 (6.2) | 1438 (11.0) |
| Nr. of negative tests<sup>4</sup>, median [IQR] | 1 [0-2] | 2 [1-4] | 2 [1-4] |
| Nr. of all-cause previous visits to primary care<sup>4</sup>, median [IQR] | 1 [1-3] | 2 [1-3] | 1 [0-3] |

All characteristics measured prior to test. <sup>1</sup> Highest achieved education level by end of 2019. <sup>2</sup> Based on counts of medical records in specialist care, January 1<sup>st</sup> 2020 to December 7<sup>th</sup> 2021. <sup>3</sup> The current vaccination status against COVID-19 before selected test date, based on records in vaccination database. <sup>4</sup> Counts of registered tests, from January 1<sup>st</sup> 2020 to December 7<sup>th</sup> 2021. <sup>5</sup> Counts of contacts with general practitioner or emergency ward, from January 1<sup>st</sup> 2020 to December 7<sup>th</sup> 2021.

**Table 3.** Person-days, numbers of failures and incidence rates per 100 000 person-days by diagnosis in primary care from 14 to up to 126 days after test date for SARS-CoV-2.

|  |  | Tested<br>negative | SARS-CoV-2 delta | SARS-CoV-2 omicron |
| --- | --- | --- | --- | --- |
| Musculoskeletal pain | Person-days | 6 327 781 | 1 404 263 | 784 982 |
|  | Failures | 5968 | 1420 | 598 |
|  | Incidence rate | 94 | 101 | 76 |
|  | (95% CI) | 92-97 | 96-107 | 70-83 |
| Fatigue | Person-days | 6 398 683 | 1 419 888 | 792 618 |
|  | Failures | 2214 | 891 | 423 |
|  | Incidence rate | 35 | 63 | 53 |
|  | (95% CI) | 33-36 | 59-67 | 49-59 |
| Cough | Person-days | 6 420 103 | 1 427 995 | 795 635 |
|  | Failures | 1043 | 298 | 128 |
|  | Incidence rate | 16 | 21 | 16 |
|  | (95% CI) | 15-17 | 19-23 | 14-19 |
| Heart palpitations | Person-days | 6 416 355 | 1 424 789 | 793 932 |
|  | Failures | 366 | 111 | 44 |
|  | Incidence rate | 6 | 8 | 6 |
|  | (95% CI) | 5-6 | 6-9 | 4-7 |
| Shortness of breath | Person-days | 6 418 779 | 1 424 687 | 794 669 |
|  | Failures | 368 | 233 | 90 |
|  | Incidence rate | 6 | 16 | 11 |
|  | (95% CI) | 5-6 | 14-19 | 9-14 |
| Anxiety/depression | Person-days | 6 390 747 | 1 419 261 | 791 621 |
|  | Failures | 2546 | 592 | 260 |
|  | Incidence rate | 40 | 42 | 33 |
|  | (95% CI) | 38-41 | 38-45 | 29-37 |
| Brain fog | Person-days | 7 146 038 | 1 587 723 | 885 166 |
|  | Failures | 143 | 41 | 18 |
|  | Incidence rate | 2 | 3 | 2 |
|  | (95% CI) | 2-3 | 2-4 | 1-4 |

Failures represent the first medical record with the diagnoses in question, from 14 to up to 126 days after the test date.

### Risk of post-covid complaints, from 14 to 126 days after test date

There was a 20% increased rate of post-covid fatigue, and 40-70% increased rate of post-covid shortness of breath, both for persons with the omicron variant and for persons with the delta variant, when compared to persons testing negative (Figure 1) (fatigue: HR=1.21 (CI :1.10-1.33) and HR=1.26 (CI: 1.17-1.35), shortness of breath: HR=1.43, CI, 1.14-1.80 and HR=1.70, CI, 1.46-1.98, respectively). Persons with omicron had similar rates of musculoskeletal pain, cough, heart palpitations, anxiety/depression, as persons with delta (Figure 2), and as persons testing negative (Figure 1).

**Figure 1.**
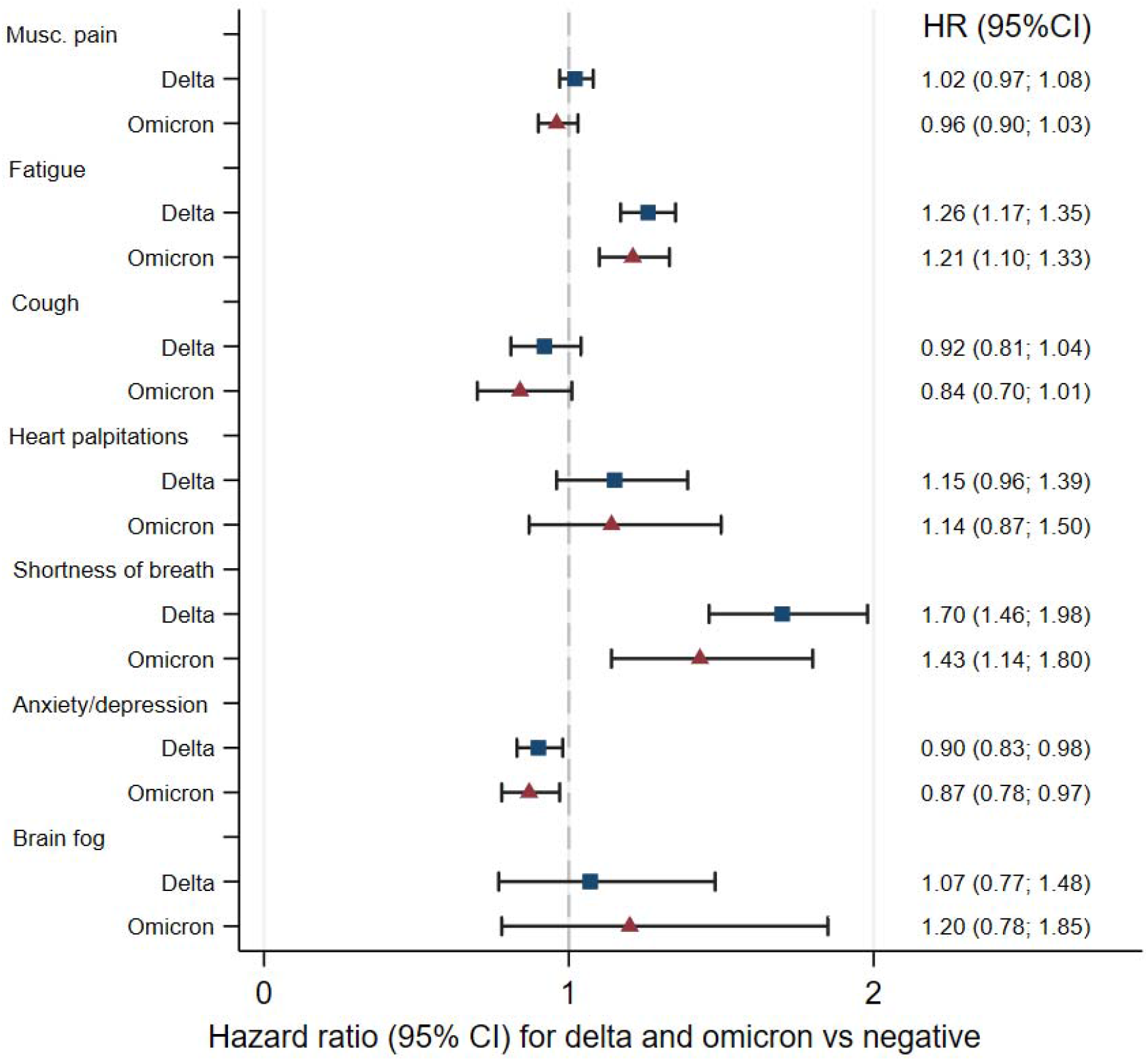
Risks of complaints from 14 to up to 126 days after SARS-CoV-2 infection with the omicron variant and after infection with the delta variant, adjusted for age, sex, education, comorbidities, test and care activity and vaccination. Reference category: persons testing negative (dashed vertical line).

**Figure 2.**
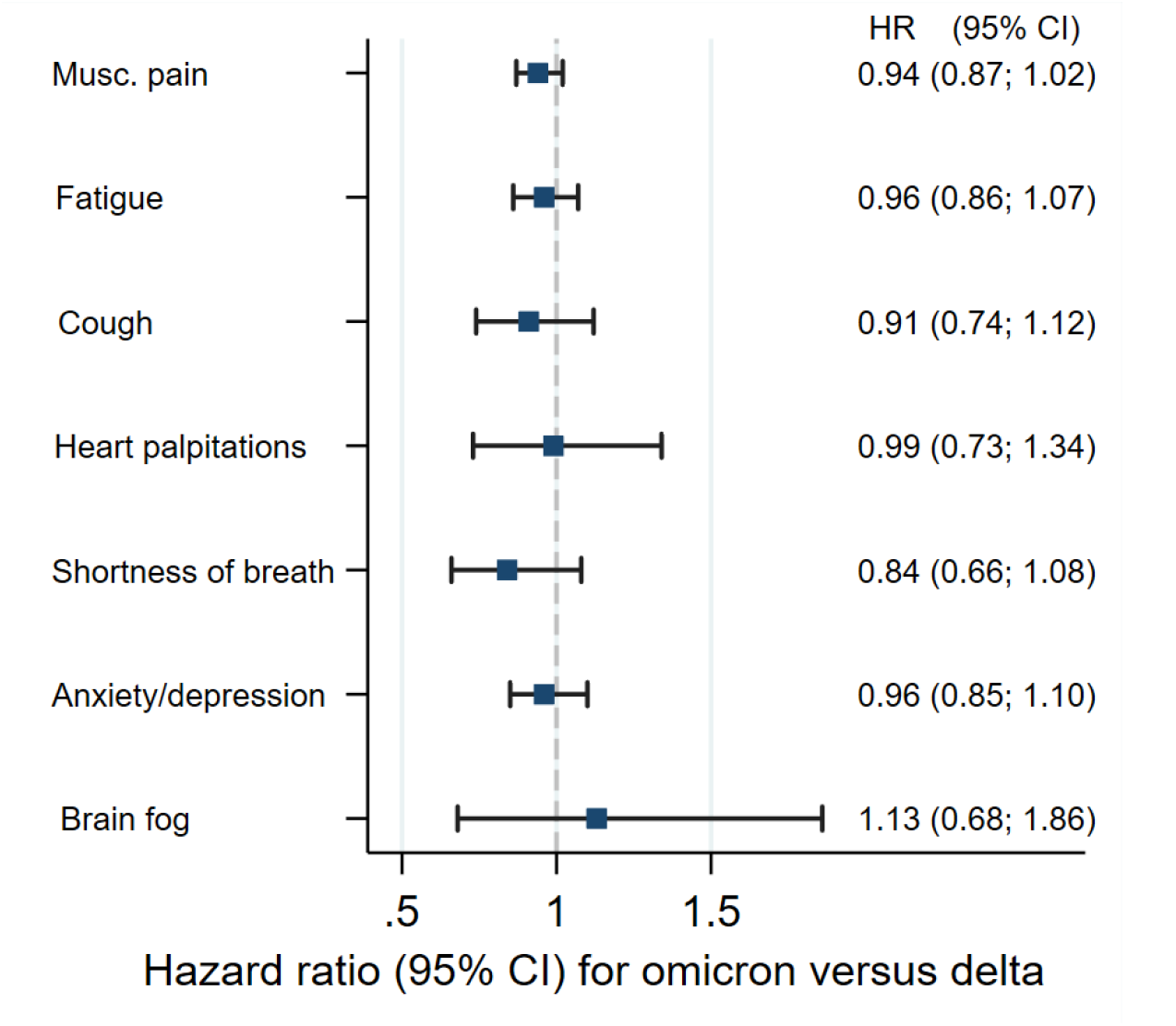
Risks of complaints from 14 to up to 126 days after SARS-CoV-2 infection with the omicron variant, adjusted for age, sex, education, comorbidities, test and care activity and vaccination. Reference category: persons with SARS-CoV-2 delta (dashed vertical line).

Crude estimates were somewhat different from adjusted estimates (S-Table 2, S-Table 3), suggesting potential confounding from sociodemographic factors or previous health and health care use. However, the estimates that were adjusted vs non-adjusted for the number of vaccine doses were rather similar, implying our findings were independent of vaccination status (S-Table 2, S-Table 3). The results were confirmed in conditional logit models matched on test week, i.e. the earlier periodical dominance of the delta variant vs the omicron variant had no or limited impact on our findings (S-Table 4, S-Table 5).

### Time-differentiated risks of post-covid complaints

When the 14 to 126 days period was separated into different clinical phases following positive test, we again observed an increased risk of fatigue and shortness of breath for the first 14 to 30 days and for the first 30 to 90 days, for persons with omicron and delta vs persons testing negative (Table 4). However, after more than 90 days, the risk of such complaints was lowered, for both variants. Whereas there was still a 15% increased rate for fatigue and 50% increased rate for shortness of breath at more than 90 days after delta infection when compared to persons testing negative, there was no such increased risk after omicron infection (HR∼1 for both outcomes, relative to test negative) (Table 4). Still, the evidence implying that infection with the omicron variant was associated with a lower risk of fatigue and shortness of breath during this later period when compared to the delta variant only was inconclusive due to wide CIs (Table 5).

**Table 4.** Estimated risks of post-covid diagnoses in primary care for different periods after testing for SARS-CoV-2, for persons with the omicron and delta variant relative to non-infected (testing negative).

| Outcome | SARS-CoV-2 status | 14 to 30 days |  | 30 to 90 days |  | 90 to 126 days |  |
| --- | --- | --- | --- | --- | --- | --- | --- |
|  |  | HR | 95% CI | HR | 95% CI | HR | 95% CI |
| Musculoskeletal pain | Non-infected | 1 |  | 1 |  | 1 |  |
|  | Delta | 1.02 | [0.92,1.13] | 1.00 | [0.94,1.06] | 1.06 | [0.98,1.16] |
|  | Omicron | 0.77 | [0.66,0.89] | 0.92 | [0.84,1.00] | 0.98 | [0.85,1.13] |
| Fatigue | Non-infected | 1 |  | 1 |  | 1 |  |
|  | Delta | 1.85 | [1.61,2.13] | 1.35 | [1.24,1.47] | 1.15 | [1.00,1.31] |
|  | Omicron | 1.50 | [1.24,1.82] | 1.25 | [1.12,1.40] | 1.00 | [0.81,1.25] |
| Cough | Non-infected | 1 |  | 1 |  | 1 |  |
|  | Delta | 1.21 | [0.96,1.52] | 0.91 | [0.78,1.07] | 0.71 | [0.54,0.92] |
|  | Omicron | 1.28 | [0.94,1.74] | 0.75 | [0.59,0.95] | 0.77 | [0.52,1.15] |
| Heart palpitations | Non-infected | 1 |  | 1 |  | 1 |  |
|  | Delta | 1.18 | [0.80,1.75] | 1.30 | [1.04,1.63] | 1.00 | [0.69,1.46] |
|  | Omicron | 0.88 | [0.50,1.52] | 1.04 | [0.74,1.47] | 1.33 | [0.77,2.29] |
| Shortness of breath | Non-infected | 1 |  | 1 |  | 1 |  |
|  | Delta | 3.54 | [2.65,4.71] | 2.02 | [1.69,2.43] | 1.57 | [1.14,2.16] |
|  | Omicron | 2.48 | [1.67,3.69] | 1.32 | [0.99,1.76] | 0.87 | [0.46,1.65] |
| Anxiety/depression | Non-infected | 1 |  | 1 |  | 1 |  |
|  | Delta | 0.87 | [0.75,1.01] | 0.89 | [0.81,0.97] | 0.92 | [0.81,1.04] |
|  | Omicron | 0.79 | [0.65,0.98] | 0.80 | [0.70,0.91] | 0.90 | [0.74,1.10] |
| Brain fog | Non-infected | 1 |  | 1 |  | 1 |  |
|  | Delta | 0.86 | [0.44,1.71] | 1.09 | [0.75,1.59] | 0.69 | [0.33,1.42] |
|  | Omicron | 0.53 | [0.19,1.54] | 1.31 | [0.80,2.12] | 1.12 | [0.46,2.73] |

\*Adjusted for age, sex, education status, country of birth, the number of comorbidities, the number of negative tests, the number of primary care visits and the number of vaccine doses prior to the selected test date. Stratified on calendar week of testing.

**Table 5.** Estimated risks of post-covid diagnoses in primary care for different time periods after testing for SARS-CoV-2, for persons with the omicron variant vs the delta variant.

| Outcome | SARS-CoV-2 status | 14 to 30 days |  | 30 to 90 days |  | 90 to 126 days |  |
| --- | --- | --- | --- | --- | --- | --- | --- |
|  |  | HR | 95% CI | HR | 95% CI | HR | 95% CI |
| Musculoskeletal pain | Delta | 1 |  | 1 |  | 1 |  |
|  | Omicron | 0.76 | [0.64,0.89] | 0.92 | [0.83,1.01] | 0.91 | [0.78,1.07] |
| Fatigue | Delta | 1 |  | 1 |  | 1 |  |
|  | Omicron | 0.81 | [0.66,1.00] | 0.93 | [0.82,1.05] | 0.94 | [0.74,1.19] |
| Cough | Delta | 1 |  | 1 |  | 1 |  |
|  | Omicron | 1.06 | [0.74,1.50] | 0.82 | [0.63,1.08] | 1.1 | [0.70,1.73] |
| Heart palpitations | Delta | 1 |  | 1 |  | 1 |  |
|  | Omicron | 0.74 | [0.40,1.38] | 0.8 | [0.54,1.17] | 1.35 | [0.71,2.57] |
| Shortness of breath | Delta | 1 |  | 1 |  | 1 |  |
|  | Omicron | 0.7 | [0.47,1.06] | 0.65 | [0.48,0.89] | 0.57 | [0.28,1.16] |
| Anxiety/depression | Delta | 1 |  | 1 |  | 1 |  |
|  | Omicron | 0.91 | [0.72,1.15] | 0.9 | [0.78,1.04] | 0.97 | [0.78,1.22] |
| Brain fog | Delta | 1 |  | 1 |  | 1 |  |
|  | Omicron | 0.62 | [0.19,2.02] | 1.2 | [0.68,2.10] | 1.38 | [0.44,4.33] |

\*Adjusted for age, sex, education status, country of birth, the number of comorbidities, the number of negative tests, the number of primary care visits and the number of vaccine doses prior to the selected test date. Stratified on calendar week of testing.

Results were indicative of similar findings when studied as crude rates (S-Table 6-8) as well as in sensitivity analyses using the group of untested persons (N=1 373 092, mean (SD) age 46.5 (14.9), women n=609 189 (44%), primary school n=315 790 (23%), born in Norway n=1 054 690 (77%), 2 vaccine doses n=1 120 219 (82%), median (IQR) previous care visits 0 (0-2)) as comparison group (S-Table 9). However, both infections with the delta and omicron variants were associated with an increased risk of cough for periods 14 to 30 days when compared to untested (S-Table 9), which was not observed when compared to persons testing negative in this period (Table 4).

## Discussion

In this population-based prospective cohort study, we found that persons with omicron and delta both had approximately 20% increased risk of a healthcare diagnosis of post-covid fatigue and up to 40-70% increased risk of post-covid shortness of breath, compared to persons testing negative, for up to 126 days after being tested. The risk of complaints was the highest in the acute phase (14 to 30 days) and decreased for both variants in the sub-acute (30 to 90 days) and assumed chronic post-covid condition (here: 90 to 126 days) phases. Persons with either of the two variants had similar, and no increased rates of musculoskeletal pain, cough, heart palpitations, anxiety/depression and brain fog in any of the post-covid clinical phases.

### Comparison to previous studies

This is to our awareness the first study to provide estimates of sequelae of the omicron variant. As such, it sheds new and important light on the growing body of evidence suggesting that omicron leads to milder acute disease and fewer hospitalizations than delta^4^. We found no study of post-covid medical records in primary care for an effective comparison of findings to previous studies on other SARS-CoV-2 variants. However, Lund et al. reported that non-hospitalized persons with COVID-19 (unknown variant but assumed to be the earliest and original one) had a doubled risk of having a hospital diagnosis of shortness of breath for up to 6 months after the initial disease compared to a matched non-infected cohort (risk ratios of 2.00, 95% CI=1.62– 2.48)^1^.

These estimates are of slightly higher magnitude as observed for our full-period (∼4 months), i.e. when studying outpatient diagnoses of shortness of breath for omicron and delta (HR1.43, 95% CI=1.14-1.80 and HR=1.70, 95% CI=1.46-1.98, respectively). In contrast, the two studies’ estimates of post-covid fatigue are differing (risk ratio=0.97, 95%CI = 0.60-1.59 in Lund et al., and HR=1.21, 95% CI=1.10-1.33 for omicron and HR=1.26, 95% CI=1.17-1.35 for delta in the present study). Potential explanations for the higher risk of fatigue (at least for up to 90 days after test date for omicron) in the current than in previous studies^1^ may be the care level in which the complaints are seen. General practitioners may be less likely to refer to specialist care when the main complaint is fatigue than when the main complaint is shortness of breath. We did not aim to study post-covid outcomes in specialist care in the current study, as our previous studies using similar data sources suggested no impact of mild SARS-CoV-2 infection on the hospital care services^2^. However, we believe the similar findings for the delta variant as seen for the earliest variants^1^ (two variants with an assumed similar disease severity) together with the similar findings for omicron and delta in the current study support the interpretation that the prevalence of post-covid complaints is similar after omicron than after earlier variants. As such, two large Nordic register-based studies together indicate that mainly persistent shortness of breath and to a lower extent fatigue pose a burden on the post-covid healthcare services in the omicron strain, yet that this burden will cease when 90 days have passed after infection. In neither of the studies were the rates of the other potential post-covid complaints increased.

### Relevance to public health, clinic, and future research

Overall, our findings suggest that the included post-covid complaints exist to a similar extent after omicron as after delta. However, the risk of fatigue and shortness of breath was greatly lowered over time for both variants, shedding new light on the risk of the post-covid condition. which was recently defined by the World Health Organization^12^ (persistent complaints, typically fatigue and shortness of breath, with unknown cause still present at 3 months from the onset). We observed rather minor increased risks of these complaints at 3 months or more after onset when compared to persons testing negative (Table 4), and no increased risk when compared to untested persons (S-Table 9). Further, given no observed differences between omicron and delta in direct comparisons of time-differentiated risks (Table 5, S-Table 9), our findings suggest that both the omicron and delta variants will lead to a temporarily increased number of care visits and rather similar burdens on the healthcare services in the long run. More specifically, the high number of cases of symptomatic SARS-CoV-2 omicron, even in vaccinated individuals^7^, suggests that we may expect a substantial increase in the number of patients with fatigue and shortness of breath a few weeks after a large wave of transmission.

Our findings may have some important combined clinical and public health messages. First, we provide novel insights into disease etiology of post-omicron, demonstrating the need to further study the onset, duration and severity of post-covid complaints following the omicron variant, e.g. using patient-reported or clinical data. As such, omicron might be hypothesized to lead to mild post-covid disease in the long run, but not in the short run or sub-acute phase. Second, we provide data material to be used by public health workers, policy makers and clinicians, for example for cost estimation and prioritizing resources in primary care in times or regions where many are infected with omicron simultaneously.

### Strengths and limitations

Strengths of our study include the use of sequenced data allowing comparison of omicron vs delta during the same calendar period when the two variants had the largest overlap, combined with health care register data with no attrition. Further, equal access to SARS-CoV-2 testing at no cost for the individuals as well as a universal tax-funded health care system, improve generalizability of findings to other countries.

A limitation of our study is that we could not include antigen or home tests as they were not registered. Polymerase chain reaction testing was however mandatory for everyone with a positive antigen test during our study period. Moreover, all parcitipants in our study had a PCR test in a period characterized by great uncertainty regarding the severity of the new SARS-CoV-2 omicron variant. It is possible that our population consisted of particularly health-conscious persons who were highly prone to get tested and who were more prone to seek medical care after knowing they had been ill. We believe our methodological approach ensuring comparison of persons who were tested in the same calendar week limits this potential bias arising due to anxiety. Further, we have previously reported that register-based studies comparing persons with positive test with persons with negative test likely will lead to an underestimation of post-covid outcomes’ prevalence, as persons testing themselves may represent a particularly health-conscious sample of the population^3^. Indeed, when we repeated the analyses using a group of untested persons as a comparison group, the estimated hazard ratios were higher, at least for the acute phase (S-Table 9). However, the untested group may be hypothesized to consist of persons who are particularly unconscious about their health, potentially leading to overestimation of post-covid outcomes’ prevalence^3^. Finally, a limitation of our study was the few observations of brain fog and estimates should be interpreted with care. Also, some other outcomes in the period 90 to 126 days had low absolute counts, leading to wide CIs and potentially inefficient confounding adjustment.

In conclusion, we found that SARS-CoV-2 omicron and delta infection are associated with similarly increased risks of fatigue and shortness of breath when compared to non-infected. The omicron variant will likely lead to a temporarily increased burden on healthcare services.

## Data Availability

The study method and statistical analyses are all described in detail in the Methods chapter and throughout the paper. Individual-level data of patients included in this paper after de-identification are considered sensitive and will not be shared. However, the individual-level data in the registries compiled in Beredt C19 are accessible to authorized researchers after ethical approval and application to "helsedata.no/en" administered by the Norwegian Directorate of eHealth. Data requests may be sent to ".

## Acknowledgements

We thank the Norwegian Directorate of Health, particularly Olav Isak Sjøflot and his Department of Health Registries for their cooperation in establishing the emergency preparedness register, and Gutorm Høgåsen and Anja Elsrud Schou Lindman for their invaluable work on the register. The interpretation and reporting of the data are the sole responsibility of the authors, and no endorsement by the register is intended or should be inferred. We also thank staff at the Norwegian Institute of Public Health who have been part of the outbreak investigation and response team.

## Author contribution

Karin Magnusson had full access to all of the data in the study and take responsibility for the integrity of the data and the accuracy of the data analysis.

*Concept and design:* Magnusson, Kinge

*Acquisition, analysis, or interpretation of data:* All authors.

*Drafting of the manuscript:* Magnusson

*Critical revision of the manuscript for important intellectual content:* All authors

*Statistical analysis:* Magnusson, Turkiewicz, Kinge

*Obtained funding:* Englund

*Administrative, technical, or material support:* N/A

*Supervision:* N/A

All authors contributed in drafting the article or critically revising it for important intellectual content. All authors gave final approval for the version to be submitted.

## Supplementary material for the paper

**Supplementary Table 1.**
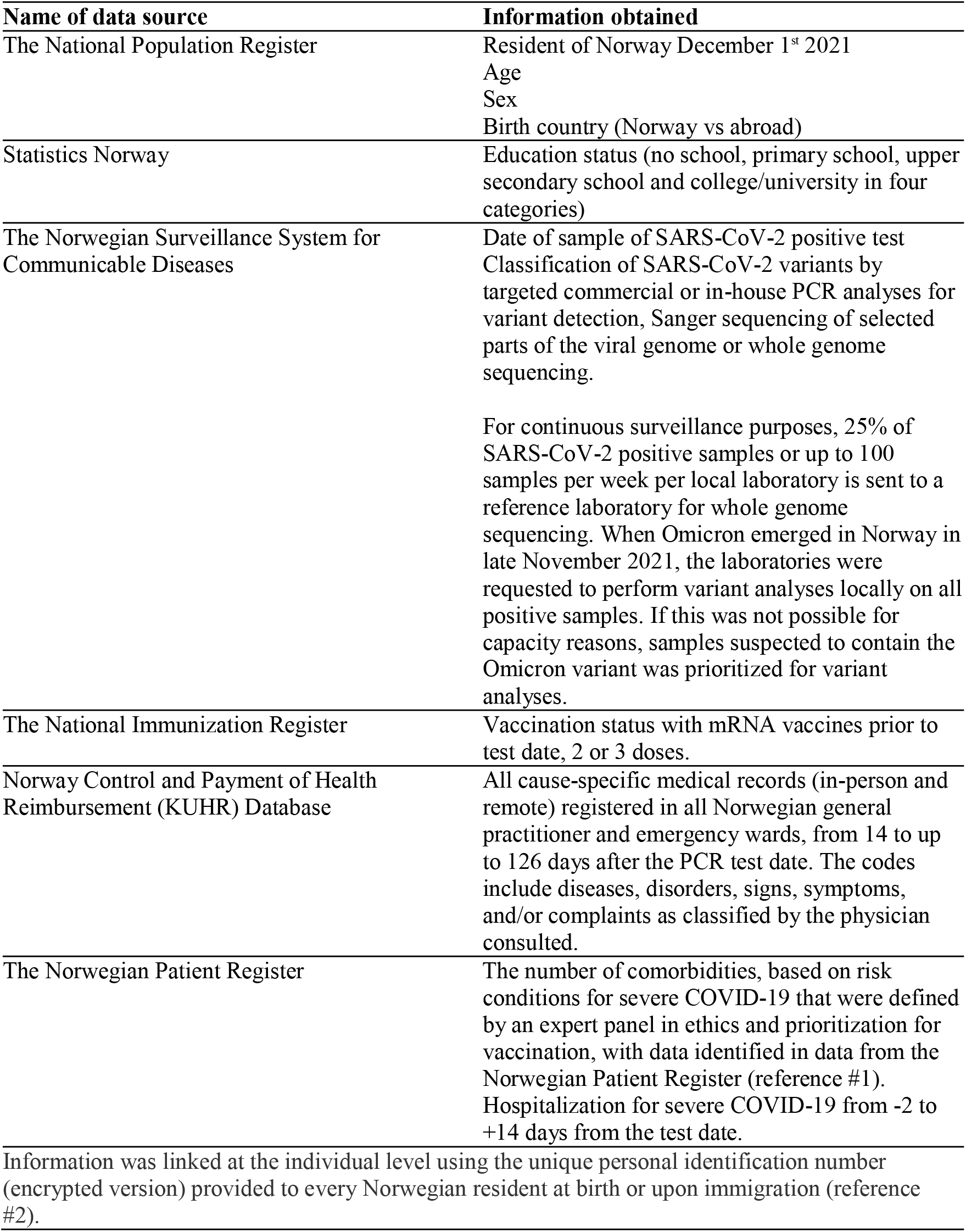
Data sources in Beredt C19 used in this study and information obtained from each source.

| **Name of data source** | **Information obtained** |
| --- | --- |
| The National Population Register | Resident of Norway December 1<sup>st</sup> 2021<br>Age<br>Sex<br>Birth country (Norway vs abroad) |
| Statistics Norway | Education status (no school, primary school, upper secondary school and college/university in four categories) |
| The Norwegian Surveillance System for Communicable Diseases | Date of sample of SARS-CoV-2 positive test<br>Classification of SARS-CoV-2 variants by targeted commercial or in-house PCR analyses for variant detection, Sanger sequencing of selected parts of the viral genome or whole genome sequencing.<br>For continuous surveillance purposes, 25% of SARS-CoV-2 positive samples or up to 100 samples per week per local laboratory is sent to a reference laboratory for whole genome sequencing. When Omicron emerged in Norway in late November 2021, the laboratories were requested to perform variant analyses locally on all positive samples. If this was not possible for capacity reasons, samples suspected to contain the Omicron variant was prioritized for variant analyses. |
| The National Immunization Register | Vaccination status with mRNA vaccines prior to test date, 2 or 3 doses. |
| Norway Control and Payment of Health Reimbursement (KUHR) Database | All cause-specific medical records (in-person and remote) registered in all Norwegian general practitioner and emergency wards, from 14 to up to 126 days after the PCR test date. The codes include diseases, disorders, signs, symptoms, and/or complaints as classified by the physician consulted. |
| The Norwegian Patient Register | The number of comorbidities, based on risk conditions for severe COVID-19 that were defined by an expert panel in ethics and prioritization for vaccination, with data identified in data from the Norwegian Patient Register (reference #1).<br>Hospitalization for severe COVID-19 from -2 to +14 days from the test date. |
| Information was linked at the individual level using the unique personal identification number (encrypted version) provided to every Norwegian resident at birth or upon immigration (reference #2). |  |

**Supplementary Figure 1.**
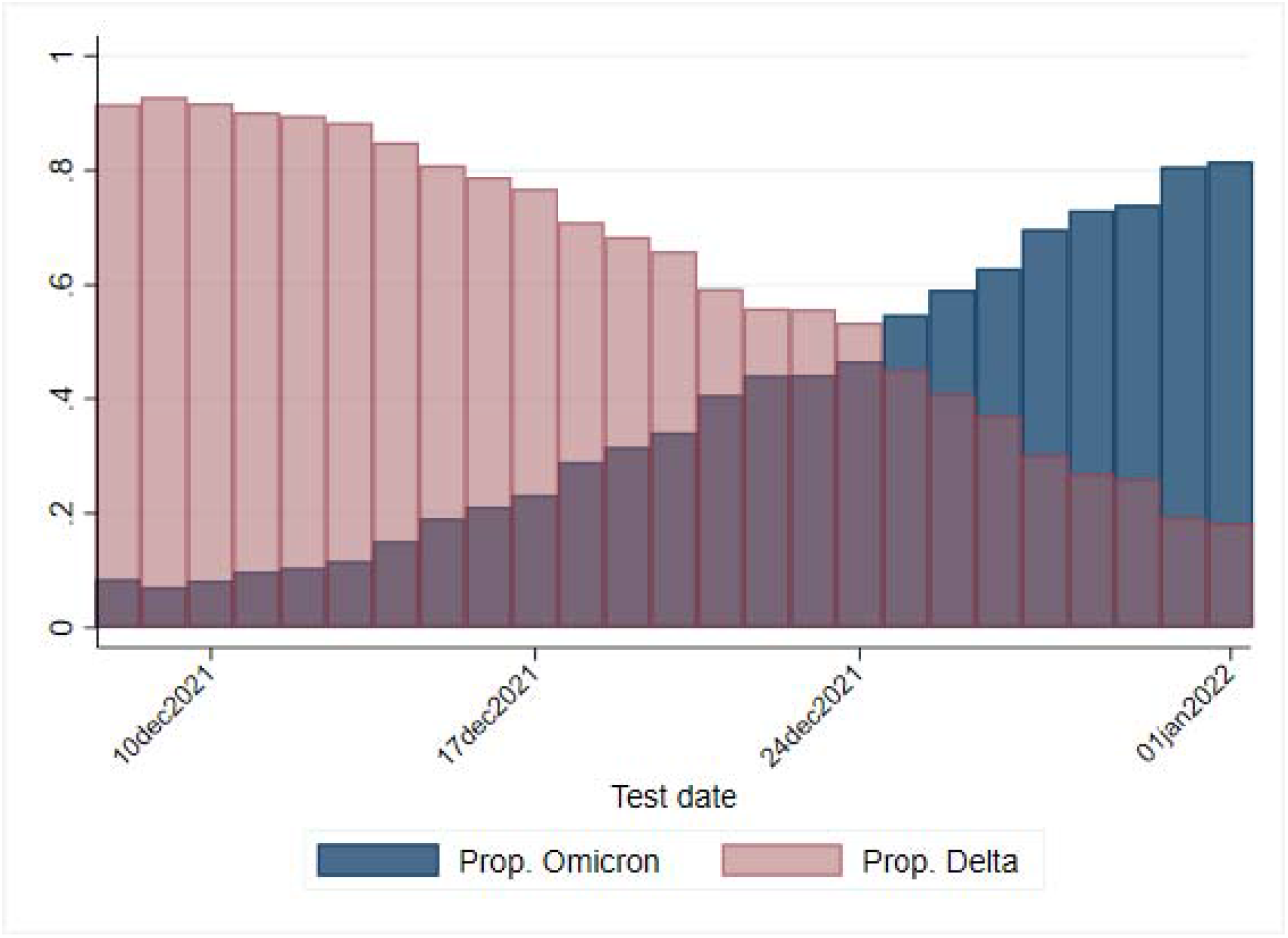
The day-by-day proportion of screened tests that were confirmed as the SARS-CoV-2 omicron or delta variants.

**Supplementary Figure 2.**
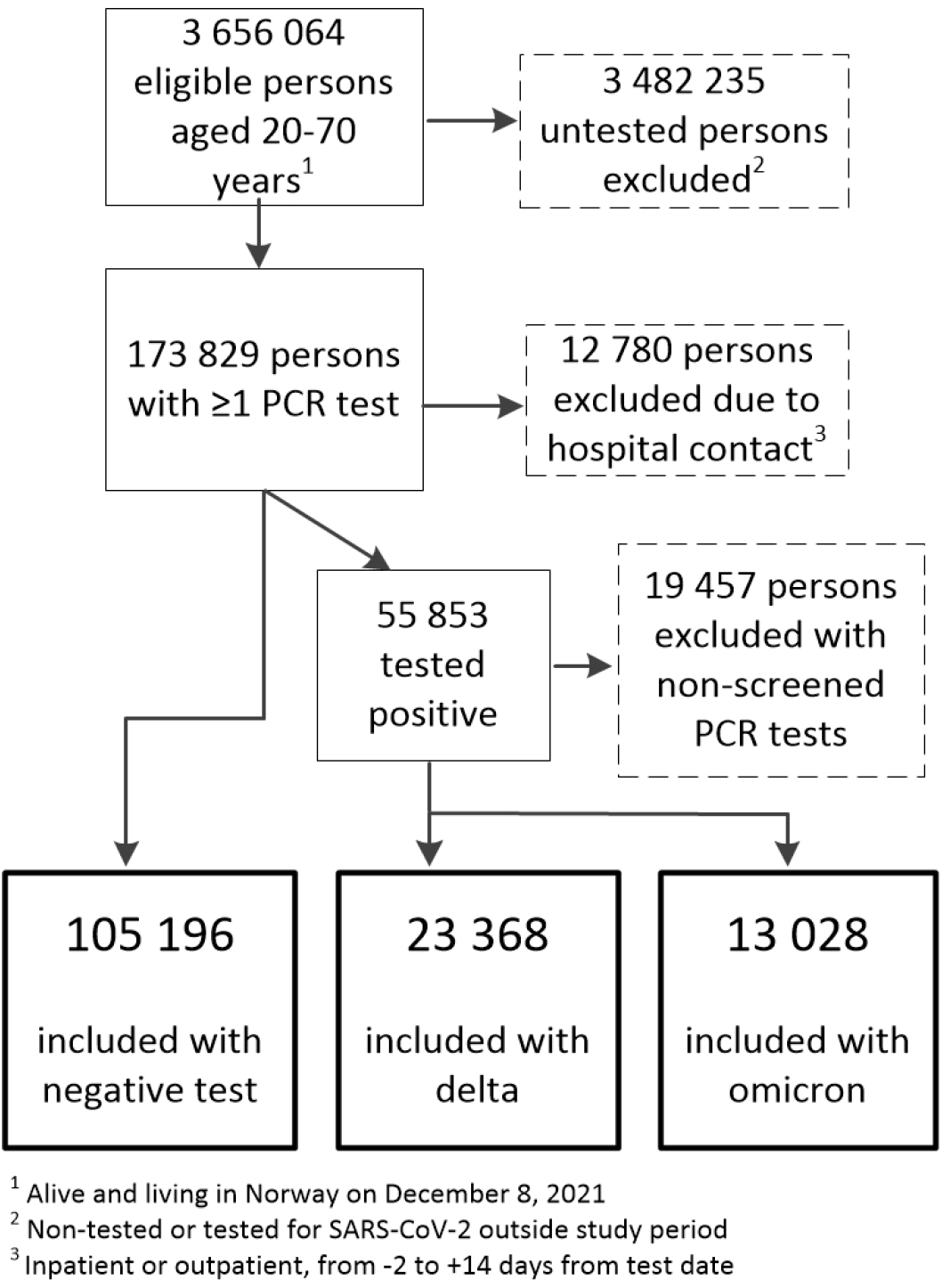
Flow chart presenting eligible, excluded and included individuals. PCR: polymerase chain reaction. Data sources described in reference #1.

**Supplementary Table 2.** Estimated hazard ratios of post-covid diagnoses in primary care from 14 to up to 126 days after testing for SARS-CoV-2, for persons with the omicron variant or delta variant, both compared to persons testing negative.

| Outcome | SARS-CoV-2 status | Crude | 95% CI | Adjusted* | 95% CI* | Adjusted* +<br>adjusted for<br>vaccination<br>status | 95% CI |
| --- | --- | --- | --- | --- | --- | --- | --- |
| Musculoskeletal pain | Tested negative | 1 |  | 1 |  | 1 |  |
|  | Delta | 1.08 | [1.03,1.13] | 1.02 | [0.97,1.07] | 1.02 | [0.97,1.08] |
|  | Omicron | 0.92 | [0.86,0.98] | 0.97 | [0.90,1.04] | 0.96 | [0.90,1.03] |
| Fatigue | Tested negative | 1 |  | 1 |  | 1 |  |
|  | Delta | 1.41 | [1.32,1.51] | 1.28 | [1.20,1.37] | 1.26 | [1.17,1.35] |
|  | Omicron | 1.37 | [1.25,1.50] | 1.22 | [1.11,1.34] | 1.21 | [1.10,1.33] |
| Cough | Tested negative | 1 |  | 1 |  | 1 |  |
|  | Delta | 0.99 | [0.88,1.11] | 0.91 | [0.81,1.03] | 0.92 | [0.81,1.04] |
|  | Omicron | 0.85 | [0.71,1.02] | 0.84 | [0.70,1.01] | 0.84 | [0.70,1.01] |
| Heart palpitations | Tested negative | 1 |  | 1 |  | 1 |  |
|  | Delta | 1.21 | [1.01,1.44] | 1.13 | [0.94,1.36] | 1.15 | [0.96,1.39] |
|  | Omicron | 1.16 | [0.89,1.52] | 1.12 | [0.85,1.47] | 1.14 | [0.87,1.50] |
| Shortness of breath | Tested negative | 1 |  | 1 |  | 1 |  |
|  | Delta | 1.79 | [1.55,2.07] | 1.71 | [1.47,1.99] | 1.7 | [1.46,1.98] |
|  | Omicron | 1.42 | [1.13,1.77] | 1.44 | [1.15,1.81] | 1.43 | [1.14,1.80] |
| Anxiety/depression | Tested negative | 1 |  | 1 |  | 1 |  |
|  | Delta | 1.06 | [0.98,1.14] | 0.93 | [0.86,1.01] | 0.9 | [0.83,0.98] |
|  | Omicron | 0.96 | [0.86,1.07] | 0.88 | [0.79,0.99] | 0.87 | [0.78,0.97] |
| Brain fog | Tested negative | 1 |  | 1 |  | 1 |  |
|  | Delta | 1.17 | [0.86,1.59] | 1.04 | [0.75,1.43] | 1.07 | [0.77,1.48] |
|  | Omicron | 1.29 | [0.85,1.98] | 1.19 | [0.78,1.84] | 1.2 | [0.78,1.85] |

\*Adjusted for age, sex, education status, country of birth, the number of comorbidities, the number of negative test prior to the selected test date and the number of primary care visits prior to the selected test date. Stratified on calendar week of testing.

**Supplementary Table 3.** Estimated hazard ratios of post-covid diagnoses in primary care from 14 to up to 126 days after testing for SARS-CoV-2, for persons with the omicron variant vs the delta variant.

| Outcome | SARS-CoV-2 status | Crude | 95% CI | Adjusted* | 95% CI* | Adjusted* +<br>adjusted for<br>vaccination<br>status | 95% CI |
| --- | --- | --- | --- | --- | --- | --- | --- |
| Musculoskeletal pain | Delta | 1 |  | 1 |  | 1 |  |
|  | Omicron | 0.85 | [0.79,0.92] | 0.95 | [0.88,1.03] | 0.94 | [0.87,1.02] |
| Fatigue | Delta | 1 |  | 1 |  | 1 |  |
|  | Omicron | 0.97 | [0.87,1.08] | 0.95 | [0.85,1.06] | 0.96 | [0.86,1.07] |
| Cough | Delta | 1 |  | 1 |  | 1 |  |
|  | Omicron | 0.86 | [0.70,1.05] | 0.92 | [0.75,1.13] | 0.91 | [0.74,1.12] |
| Heart palpitations | Delta | 1 |  | 1 |  | 1 |  |
|  | Omicron | 0.96 | [0.71,1.30] | 0.99 | [0.73,1.34] | 0.99 | [0.73,1.34] |
| Shortness of breath | Delta | 1 |  | 1 |  | 1 |  |
|  | Omicron | 0.79 | [0.62,1.01] | 0.84 | [0.66,1.07] | 0.84 | [0.66,1.08] |
| Anxiety/depression | Delta | 1 |  | 1 |  | 1 |  |
|  | Omicron | 0.91 | [0.80,1.03] | 0.94 | [0.83,1.07] | 0.96 | [0.85,1.10] |
| Brain fog | Delta | 1 |  | 1 |  | 1 |  |
|  | Omicron | 1.11 | [0.68,1.81] | 1.15 | [0.70,1.90] | 1.13 | [0.68,1.86] |

\*Adjusted for age, sex, education status, country of birth, the number of comorbidities, the number of negative tests prior to the selected test date and the number of primary care visits prior to the selected test date. Stratified on calendar week of testing.

**Supplementary Table 4.** Estimated odds ratios of post-covid diagnoses in primary care from 14 to up to 126 days after testing for SARS-CoV-2, for persons with the omicron variant or delta variant, both compared to persons testing negative.

| Outcome | SARS-CoV-2 status | Crude | 95% CI | Adjusted* | 95% CI* | Adjusted* + adjusted for vaccination status | 95% CI |
| --- | --- | --- | --- | --- | --- | --- | --- |
| Musculoskeletal pain | Tested negative | 1 |  | 1 |  | 1 |  |
|  | Delta | 1.09 | [1.03,1.15] | 1.02 | [0.96,1.09] | 1.03 | [0.96,1.09] |
|  | Omicron | 0.89 | [0.82,0.97] | 0.95 | [0.87,1.04] | 0.95 | [0.86,1.04] |
| Fatigue | Tested negative | 1 |  | 1 |  | 1 |  |
|  | Delta | 1.49 | [1.37,1.62] | 1.38 | [1.26,1.51] | 1.37 | [1.25,1.50] |
|  | Omicron | 1.32 | [1.18,1.48] | 1.18 | [1.05,1.34] | 1.18 | [1.05,1.33] |
| Cough | Tested negative | 1 |  | 1 |  | 1 |  |
|  | Delta | 0.99 | [0.85,1.14] | 0.92 | [0.79,1.08] | 0.93 | [0.80,1.09] |
|  | Omicron | 0.84 | [0.68,1.04] | 0.85 | [0.69,1.06] | 0.85 | [0.68,1.06] |
| Heart palpitations | Tested negative | 1 |  | 1 |  | 1 |  |
|  | Delta | 1.35 | [1.08,1.68] | 1.14 | [0.90,1.45] | 1.22 | [0.95,1.57] |
|  | Omicron | 1.33 | [0.95,1.87] | 1.28 | [0.89,1.83] | 1.32 | [0.92,1.90] |
| Shortness of breath | Tested negative | 1 |  | 1 |  | 1 |  |
|  | Delta | 1.9 | [1.58,2.29] | 1.9 | [1.55,2.33] | 1.91 | [1.55,2.35] |
|  | Omicron | 1.65 | [1.24,2.20] | 1.64 | [1.21,2.24] | 1.63 | [1.20,2.22] |
| Anxiety/depression | Tested negative | 1 |  | 1 |  | 1 |  |
|  | Delta | 1.07 | [0.98,1.18] | 0.95 | [0.85,1.05] | 0.92 | [0.83,1.02] |
|  | Omicron | 1.03 | [0.90,1.18] | 0.94 | [0.81,1.08] | 0.93 | [0.80,1.07] |
| Brain fog | Tested negative | 1 |  | 1 |  | 1 |  |
|  | Delta | 1.15 | [0.78,1.70] | 0.97 | [0.62,1.51] | 0.99 | [0.63,1.55] |
|  | Omicron | 1.42 | [0.83,2.43] | 1.32 | [0.73,2.37] | 1.33 | [0.74,2.39] |

\*Adjusted for age, sex, education status, country of birth, the number of comorbidities, the number of negative tests and the number of primary care visits prior to the selected test date. Estimates are from conditional logit models with matching of cases (persons with the outcome in question) and controls (persons without the outcome in question, two controls per case) on their calendar week of testing.

**Supplementary Table 5.** Estimated odds ratios of post-covid diagnoses in primary care from 14 to up to 126 days after testing for SARS-CoV-2, for persons with the omicron variant vs the delta variant.

| Outcome | SARS-CoV-2 status | Crude | 95% CI | Adjusted* | 95% CI* | Adjusted* + adjusted for vaccination status | 95% CI |
| --- | --- | --- | --- | --- | --- | --- | --- |
| Musculoskeletal pain | Delta | 1 |  | 1 |  | 1 |  |
|  | Omicron | 0.82 | [0.75,0.91] | 0.93 | [0.84,1.03] | 0.92 | [0.83,1.02] |
| Fatigue | Delta | 1 |  | 1 |  | 1 |  |
|  | Omicron | 0.89 | [0.78,1.01] | 0.86 | [0.75,0.99] | 0.86 | [0.75,0.99] |
| Cough | Delta | 1 |  | 1 |  | 1 |  |
|  | Omicron | 0.85 | [0.67,1.09] | 0.92 | [0.72,1.19] | 0.91 | [0.71,1.18] |
| Heart palpitations | Delta | 1 |  | 1 |  | 1 |  |
|  | Omicron | 0.99 | [0.67,1.45] | 1.12 | [0.75,1.67] | 1.08 | [0.72,1.62] |
| Shortness of breath | Delta | 1 |  | 1 |  | 1 |  |
|  | Omicron | 0.87 | [0.63,1.19] | 0.87 | [0.62,1.21] | 0.86 | [0.61,1.20] |
| Anxiety/depression | Delta | 1 |  | 1 |  | 1 |  |
|  | Omicron | 0.96 | [0.82,1.12] | 0.99 | [0.84,1.17] | 1.01 | [0.86,1.19] |
| Brain fog | Delta | 1 |  | 1 |  | 1 |  |
|  | Omicron | 1.24 | [0.67,2.29] | 1.35 | [0.69,2.66] | 1.34 | [0.68,2.64] |

\*Adjusted for age, sex, education status, country of birth, the number of comorbidities, the number of negative tests and the number of primary care visits prior to the selected test date. Estimates are from conditional logit models with matching of cases (persons with the outcome in question) and controls (persons without the outcome in question, two controls per case) on their calendar week of testing.

**Supplementary Table 6.** Person-days, numbers of failures and incidence rates per 100 000 person-days by diagnosis in primary care after test date for SARS-CoV-2.

|  |  | 14 to 30 days |  |  |  |
| --- | --- | --- | --- | --- | --- |
|  |  | Tested negative | SARS-CoV-2 delta | SARS-CoV-2 omicron | Untested |
| Musculoskeletal pain | Person-days | 1769930 | 392848 | 219389 | 23121680 |
|  | Failures | 2083 | 504 | 214 | 25558 |
|  | Incidence rate | 118 | 128 | 98 | 111 |
|  | (95% CI) | 113-123 | 118-140 | 85-112 | 109-112 |
| Fatigue | Person-days | 1779786 | 393847 | 219935 | 23254340 |
|  | Failures | 686 | 327 | 151 | 6699 |
|  | Incidence rate | 39 | 83 | 69 | 29 |
|  | (95% CI) | 36-42 | 74-93 | 59-81 | 28-30 |
| Cough | Person-days | 1781528 | 395370 | 220563 | 23283188 |
|  | Failures | 361 | 104 | 54 | 2011 |
|  | Incidence rate | 20 | 26 | 24 | 9 |
|  | (95% CI) | 18-22 | 22-32 | 19-32 | 8-9 |
| Heart palpitations | Person-days | 1783098 | 395845 | 220789 | 23287966 |
|  | Failures | 124 | 36 | 16 | 1329 |
|  | Incidence rate | 7 | 9 | 7 | 6 |
|  | (95% CI) | 6-8 | 7-13 | 4-12 | 5-6 |
| Shortness of breath | Person-days | 1783293 | 395497 | 220720 | 23289836 |
|  | Failures | 116 | 97 | 38 | 1084 |
|  | Incidence rate | 7 | 25 | 17 | 5 |
|  | (95% CI) | 5-7 | 20-30 | 13-24 | 4-5 |
| Anxiety/depression | Person-days | 1777239 | 394632 | 220239 | 23218884 |
|  | Failures | 1023 | 228 | 113 | 11614 |
|  | Incidence rate | 58 | 58 | 51 | 50 |
|  | (95% CI) | 54-61 | 51-66 | 43-62 | 49-51 |
| Brain fog | Person-days | 1783540 | 395924 | 220900 | 23293022 |
|  | Failures | 54 | 11 | 4 | 489 |
|  | Incidence rate | 3 | 3 | 2 | 2 |
|  | (95% CI) | 2-4 | 2-5 | 1-5 | 2-2 |

Failures represent the first medical record with the diagnoses in question for the period in question.

**Supplementary Table 7.** Person-days, numbers of failures and incidence rates per 100 000 person-days by diagnosis in primary care after test date for SARS-CoV-2.

|  |  | 30 to 90 days |  |  |  |
| --- | --- | --- | --- | --- | --- |
|  |  | Tested<br>negative | SARS-CoV-2<br>delta | SARS-CoV-2<br>omicron | Untested |
| Musculoskeletal<br>pain | Person-days | 6018229 | 1333881 | 748214 | 78883560 |
|  | Failures | 6108 | 1452 | 676 | 69545 |
|  | Incidence<br>rate | 101 | 109 | 90 | 88 |
|  | (95%CI) | 99-104 | 103-115 | 84-97 | 88-89 |
| Fatigue | Person-days | 6141909 | 1355128 | 756445 | 80436248 |
|  | Failures | 2329 | 798 | 411 | 20659 |
|  | Incidence<br>rate | 38 | 59 | 54 | 26 |
|  | (95%CI) | 36-40 | 55-63 | 49-60 | 25-26 |
| Cough | Person-days | 6181430 | 1373029 | 766151 | 80835520 |
|  | Failures | 862 | 200 | 81 | 6900 |
|  | Incidence<br>rate | 14 | 15 | 11 | 9 |
|  | (95%CI) | 13-15 | 13-17 | 41518 | 8-9 |
| Heart palpitations | Person-days | 6195088 | 1375639 | 767247 | 80901136 |
|  | Failures | 358 | 110 | 40 | 4139 |
|  | Incidence<br>rate | 6 | 8 | 5 | 5 |
|  | (95%CI) | 5-6 | 7-10 | 4-7 | 5-5 |
| Shortness of breath | Person-days | 6194134 | 1372613 | 766703 | 80904248 |
|  | Failures | 399 | 196 | 61 | 4164 |
|  | Incidence<br>rate | 6 | 14 | 8 | 5 |
|  | (95%CI) | 6-7 | 12-16 | 6-10 | 5-5 |
| Anxiety/depression | Person-days | 6122842 | 1359074 | 760438 | 80126272 |
|  | Failures | 2634 | 617 | 281 | 28802 |
|  | Incidence<br>rate | 43 | 45 | 37 | 36 |
|  | (95%CI) | 41-45 | 42-49 | 33-42 | 36-36 |
| Brain fog | Person-days | 6200338 | 1377558 | 767604 | 80972584 |
|  | Failures | 138 | 38 | 22 | 1528 |
|  | Incidence<br>rate | 2 | 3 | 3 | 2 |
|  | (95%CI) | 2-3 | 2-4 | 2-4 | 2-2 |

Failures represent the first medical record with the diagnoses in question for the period in question.

**Supplementary Table 8.** Person-days, numbers of failures and incidence rates per 100 000 person-days by diagnosis in primary care after test date for SARS-CoV-2.

|  |  | 90 to 126 days |  |  |  |
| --- | --- | --- | --- | --- | --- |
|  |  | Tested negative | SARS-CoV-2 delta | SARS-CoV-2 omicron | Untested |
| Musculoskeletal pain | Person-days | 2618456 | 612775 | 242264 | 31842790 |
|  | Failures | 2800 | 725 | 227 | 30452 |
|  | Incidence rate | 107 | 118 | 94 | 96 |
|  | (95%CI) | 103-111 | 110-127 | 82-107 | 95-97 |
| Fatigue | Person-days | 2641401 | 619200 | 243814 | 32123476 |
|  | Failures | 996 | 300 | 97 | 8221 |
|  | Incidence rate | 38 | 48 | 40 | 26 |
|  | (95%CI) | 35-40 | 43-54 | 37-49 | 25-26 |
| Cough | Person-days | 2650440 | 622413 | 244455 | 32195304 |
|  | Failures | 421 | 72 | 29 | 2944 |
|  | Incidence rate | 16 | 12 | 12 | 9 |
|  | (95%CI) | 14-17 | 9-16 | 8-17 | 9-9 |
| Heart palpitations | Person-days | 2654091 | 622874 | 244526 | 32212196 |
|  | Failures | 159 | 36 | 16 | 1469 |
|  | Incidence rate | 6 | 6 | 7 | 5 |
|  | (95%CI) | 5-7 | 4-8 | 4-11 | 4-5 |
| Shortness of breath | Person-days | 2653976 | 622490 | 244595 | 32211106 |
|  | Failures | 149 | 58 | 11 | 1484 |
|  | Incidence rate | 6 | 9 | 4 | 5 |
|  | (95%CI) | 5-7 | 7-12 | 2-8 | 4-5 |
| Anxiety/depression | Person-days | 2638283 | 618537 | 243408 | 32055478 |
|  | Failures | 1303 | 324 | 114 | 13625 |
|  | Incidence rate | 49 | 52 | 47 | 43 |
|  | (95%CI) | 47-52 | 47-58 | 39-56 | 42-43 |
| Brain fog | Person-days | 2655355 | 623279 | 244566 | 32222228 |
|  | Failures | 58 | 9 | 6 | 604 |
|  | Incidence rate | 2 | 1 | 2 | 2 |
|  | (95%CI) | 2-3 | 2-3 | 1-5 | 2-2 |

Failures represent the first medical record with the diagnoses in question for the period in question.

**Supplementary Table 9.** Estimated risks of post-covid diagnoses in primary care for different periods after testing for SARS-CoV-2, for persons with the omicron and delta variant relative to persons who were untested.

| Outcome | SARS-CoV-2 status | 14 to 30 days |  | 30 to 90 days |  | 90 to 126 days |  |
| --- | --- | --- | --- | --- | --- | --- | --- |
|  |  | HR | 95% CI | HR | 95% CI | HR | 95% CI |
| Musculoskeletal pain | Untested | 1 |  | 1 |  | 1 |  |
|  | Delta | 0.96 | [0.81,1.13] | 0.93 | [0.84,1.03] | 0.97 | [0.83,1.12] |
|  | Omicron | 0.73 | [0.59,0.89] | 0.82 | [0.73,0.93] | 0.84 | [0.69,1.01] |
| Fatigue | Untested | 1 |  | 1 |  | 1 |  |
|  | Delta | 1.72 | [1.36,2.19] | 1.17 | [0.99,1.37] | 0.75 | [0.55,1.03] |
|  | Omicron | 1.3 | [0.99,1.70] | 1.03 | [0.86,1.24] | 0.55 | [0.38,0.78] |
| Cough | Untested | 1 |  | 1 |  | 1 |  |
|  | Delta | 2.08 | [1.37,3.15] | 1.24 | [0.92,1.66] | 0.81 | [0.50,1.31] |
|  | Omicron | 1.76 | [1.10,2.81] | 0.89 | [0.63,1.26] | 0.67 | [0.38,1.20] |
| Heart palpitations | Untested | 1 |  | 1 |  | 1 |  |
|  | Delta | 0.89 | [0.39,2.02] | 0.89 | [0.56,1.40] | 0.52 | [0.24,1.13] |
|  | Omicron | 0.69 | [0.28,1.71] | 0.6 | [0.35,1.03] | 0.47 | [0.19,1.15] |
| Shortness of breath | Untested | 1 |  | 1 |  | 1 |  |
|  | Delta | 3.52 | [2.23,5.56] | 1.94 | [1.42,2.65] | 1.15 | [0.65,2.02] |
|  | Omicron | 2.4 | [1.41,4.07] | 1.2 | [0.82,1.76] | 0.6 | [0.28,1.32] |
| Anxiety/depression | Untested | 1 |  | 1 |  | 1 |  |
|  | Delta | 0.77 | [0.60,0.98] | 0.7 | [0.59,0.82] | 0.72 | [0.57,0.90] |
|  | Omicron | 0.72 | [0.54,0.97] | 0.61 | [0.50,0.74] | 0.66 | [0.50,0.87] |
| Brain fog | Untested | 1 |  | 1 |  | 1 |  |
|  | Delta | 0.68 | [0.17,2.80] | 0.58 | [0.26,1.32] | 0.15 | [0.020,1.08] |
|  | Omicron | 0.44 | [0.084,2.27] | 0.61 | [0.25,1.47] | 0.19 | [0.023,1.52] |

\*Adjusted for age, sex, education status, country of birth, the number of comorbidities, the number of negative tests, the number of primary care visits and the number of vaccine doses prior to the selected test date. Stratified on calendar week of testing.

## References

1. Lund LC, Hallas J, Nielsen H, et al. Post-acute effects of SARS-CoV-2 infection in individuals not requiring hospital admission: a Danish population-based cohort study. The Lancet Infectious Diseases 2021. https://doi.org/10.1016/S1473-3099(21)00211-5 PMID: 33984263

2. Skyrud KD, Hernæs KH, Telle KE, Magnusson K (2021) Impacts of mild COVID-19 on elevated use of primary and specialist health care services: A nationwide register study from Norway. PLoS ONE 16(10): e0257926. https://doi.org/10.1371/journal.pone.0257926

3. Magnusson K, Skyrud K D, Suren P, et al. Healthcare use in 700 000 children and adolescents for six months after covid-19: before and after register based cohort study BMJ 2022; 376 :e066809 doi:10.1136/bmj-2021-066809

4. Maslo C, Friedland R, Toubkin M, Laubscher A, Akaloo T, Kama B. Characteristics and Outcomes of Hospitalized Patients in South Africa During the COVID-19 Omicron Wave Compared With Previous Waves. JAMA. 2022;327(6):583–584. doi:10.1001/jama.2021.24868

5. Powel A. Hints of a long COVID wave as Omicron fades. The Harvard Gazette. https://news.harvard.edu/gazette/story/2022/02/harvard-experts-expect-new-wave-of-long-covid-cases/

6. Jørgensen SB, Nygård K, Kacelnik O, Telle K. Secondary Attack Rates for Omicron and Delta Variants of SARS-CoV-2 in Norwegian Households. JAMA. 2022 Mar 7:e223780. doi: 10.1001/jama.2022.3780. Epub ahead of print.

7. Andrews N, Stowe J, Kirsebom F, Toffa S, Rickeard T, Gallagher E, et al. Covid-19 Vaccine Effectiveness against the Omicron (B.1.1.529) Variant. N Engl J Med. 2022 Mar 2. doi: 10.1056/NEJMoa2119451. Epub ahead of print. PMID: 35249272.

8. Davis, Hannah E., Gina S. Assaf, Lisa McCorkell, Hannah Wei, Ryan J. Low, Yochai Re’em, Signe Redfield, Jared P. Austin, and Athena Akrami. “Characterizing long COVID in an international cohort: 7 months of symptoms and their impact.” EClinicalMedicine 38 (2021): 101019.

9. Stavem, Knut, Waleed Ghanima, Magnus Kringstad Olsen, Hanne Margrethe Gilboe, and Gunnar Einvik. “Persistent symptoms 1.5–6 months after COVID-19 in non-hospitalised subjects: a population-based cohort study.” Thorax 76, no. 4 (2021): 405–407.

10. Norwegian Institute of Public Health. The Norwegian Emergency Preparedness Register (BEREDT C19), 2020. <https://www.fhi.no/en/id/infectious-diseases/coronavirus/emergency-preparedness-register-for-covid-19/

11. Himmels JPW, Gomez Castaneda M, Brurberg KG, Gravningen KM. COVID-19: Long-Term Symptoms after COVID-19 [Langvarige symptomer etter covid-19. Hurtigoversikt 2021] Oslo: Norwegian Institute of Public Health, 2021.

12. Sporaland GL, Mouland G, Bratland B, Rygh E, Reiso H. Allmennlegers bruk av ICPC-diagnoser og samsvar med journalnotatene. Tidsskr Nor Legeforen 2019. doi: 10.4045/tidsskr.18.0440

13. Coronavirus immunisation programme in Norway Rational for the recommendations. 2020, Norwegian Insititute of Public Health: Oslo. <https://www.fhi.no/contentassets/9d21BQZKqdp2CV3QV5nUEsqSg1ygegLmqRygj/norwegian-ethics-advisory-report-for-corona-vaccination.pdf

14. Nalbandian, A., Sehgal, K., Gupta, A. et al. Post-acute COVID-19 syndrome. Nat Med 27, 601–615 (2021). https://doi.org/10.1038/s41591-021-01283-z

15. Soriano JB, Murthy S, Marshall JC, Relan P, Diaz JV; WHO Clinical Case Definition Working Group on Post-COVID-19 Condition. A clinical case definition of post-COVID-19 condition by a Delphi consensus. Lancet Infect Dis. 2021 Dec 21:S1473-3099(21)00703-9. doi: 10.1016/S1473-3099(21)00703-9. Epub ahead of print. PMID: 34951953; PMCID: PMC8691845. https://pubmed.ncbi.nlm.nih.gov/34951953/

## References

1) Norwegian Institute of Public Health. The Norwegian Emergency Preparedness Register (BEREDT C19), 2020. <https://www.fhi.no/en/id/infectious-diseases/coronavirus/emergency-preparedness-register-for-covid-19/

